# Explaining Deep Neural Networks for Knowledge Discovery in Electrocardiogram Analysis

**DOI:** 10.1101/2021.01.06.20248927

**Authors:** Steven A. Hicks, Jonas L. Isaksen, Vajira Thambawita, Jonas Ghouse, Gustav Ahlberg, Allan Linneberg, Niels Grarup, Inga Strümke, Christina Ellervik, Morten Salling Olesen, Torben Hansen, Claus Graff, Niels-Henrik Holstein-Rathlou, Pål Halvorsen, Mary M. Maleckar, Michael A. Riegler, Jørgen K. Kanters

## Abstract

Deep learning-based tools may annotate and interpret medical tests more quickly, consistently, and accurately than medical doctors. However, as medical doctors remain ultimately responsible for clinical decision-making, any deep learning-based prediction must necessarily be accompanied by an explanation that can be interpreted by a human. In this study, we present an approach, called ECGradCAM, which uses attention maps to explain the reasoning behind AI decision-making and how interpreting these explanations can be used to discover new medical knowledge. Attention maps are visualizations of how a deep learning network makes, which may be used in the clinic to aid diagnosis, and in research to identify novel features and characteristics of diagnostic medical tests. Here, we showcase the use of ECGradCAM attention maps using a novel deep learning model capable of measuring both amplitudes and intervals in 12-lead electrocardiograms.

## Main

Deep learning methods have the potential to become essential tools for diagnosis and analysis in medicine. However, this family of machine learning algorithms may also bring much uncertainty and confusion among the medical practitioners they aim to help because of lacking understanding of how these algorithms work. Despite the impressive results in areas like radiology^1^, dermatology^2^, and cardiology^3–5^, deep neural networks are often criticized for being difficult to explain and for providing little to no insight into why they produce a given result (the so-called “black-box phenomenon”)^6^. Since doctors are accountable for their diagnoses, a black-box approach is unacceptable^7,8^. History has shown that doctors in practice prefer simpler although inferior algorithms to their neural network-based counterparts, primarily because the simple algorithms are more interpretable^9^. Lack of insight has in some cases of machine learning led to obvious mistakes, which has been overlooked because the black box decision did not allow understanding of how the neural network operates^10,11^. A classic example came from deep learning in radiology (X-ray of thorax), where the neural networks effectively distinguished between lung cancer and pneumonia, simply by predicting the referring department from various labels in the image and not the relevant parts of the X-ray images. When the network is presented for other X-rays without similar department labels, the network fails to distinguish between lung cancer and pneumonia^1^. This study is a good example of a mistake rooted in the differences between training and test data distribution. The neural network learned data specific features that could not be generalized to unseen data from a different distribution. This simple but grave mistake could easily be discovered with an explanation of the predictions where one could easily have observed what the network actually saw as the most important feature for its predictions. Hence, it is clear that we need to understand the decisions of the neural network. In this respect, recent developments in explainable artificial intelligence (AI) have shown progress in shedding light on these so-called “black boxes”, which seems imperative if deep learning is to be implemented in clinics^12^. Generally, explanations are produced for image data and classification. In this work, we present a method that is able to obtain explanations for classification and prediction/regression tasks and non-imaging data. Specifically, we look at electrocardiogram (ECG) where AI has become a hot topic, and where interpretable and explainable results of both classification and prediction will be crucial for clinical implementation and research.

In the field of electrocardiography, an ECG records the electrical activity of the heart using ten electrodes placed on the patient’s thorax and limbs producing 12 standard ECG leads. The ECG consists of a set of voltage time-series, with several characteristic waves (see supplemental figure S1) carrying separate clinical information about the state of the heart. The timing and the amplitude of these waves contain essential information that is associated with morbidity and mortality^13–16^. The ECG is one of the cheapest and most commonly used medical procedure, and the availability of large training data sets makes the ECG well-suited for neural network analysis. While automated analysis of ECGs has been a topic of research since the early 1960s^17^, recently, we have seen an introduction of machine learning in ECG analysis. Deep learning has shown to be successful in using features that may indicate cardiac arrhythmias or other diseases^4^.

Incorporating explainability in medical decision-making has three potential advantages. First, for implementation of deep learning in the clinic, where medical decisions may be a matter of life and death, a deep learning algorithm that explains how it arrived at a particular decision allows the prevention of rare but potentially fatal mistakes. Such mistakes may be the result of shortcomings in the training of the algorithm (such as biased or narrow training data) or noisy or faulty input data leading to unexpected and extreme decisions. Second, explainability allows researchers to potentially discover insights into medical tests and diseases. Explainability may furthermore allow for the identification of novel features in the ECGs that may lead to new understanding of the disease pathophysiology and increased diagnostic capability, which at the end may save lives. Suppose a deep learning algorithm successfully predicts sudden cardiac death using ECGs from a given population. If the algorithm also is able to explain where the information is located in the ECG, we may combine medical knowledge of the ECG with that location making it possible to identify novel mechanisms of sudden cardiac death. This would potentially make it possible to identify an intervention or possible drug target to prevent untimely death. Explainability also provides a higher level of trust and transparency in a clinical setting because a doctor can understand what the algorithm bases its predictions on. This may pave the way for implementations of neural networks in clinical practice and reduce human error, resulting in a decreased number of fatalities. Furthermore, making the algorithms more interpretable may be important from a legal perspective, where one would be able to explain why a model made an incorrect decision and place responsibility.

The work presented in this paper has three primary contributions. Firstly, we show the architecture of a residual convoluted neural network, which is able to quantify intervals and amplitudes in the ECG more accurately than trained cardiologists. Secondly, we present a modified version of the GradCAM algorithm called ECGradCAM and show how the resulting attention maps can be utilized for ECG analysis to understand, interpret, and learn from neural network predictions. The third contribution are two case studies, one where we perform predictions to measure standard clinically relevant intervals and amplitudes of the ECG and one where the network and attention maps are used to identify novel features in the ECG to learn if and how the subject’s sex can be determined from an ECG. In addition to the case studies, we also present a comparison of the assessment performance between medical doctors and the proposed model of this work.

## Results

### Automatic ECG Analysis and Data Description

We define two case-studies for model evaluation: A regression study, measuring standard clinically relevant intervals and amplitudes of the ECG, and a classification study, to predict the sex from the ECG. With respect to the first case study, numerous cardiovascular diseases are diagnosed through the measurement of key intervals and amplitudes present in the ECG. We leverage this to predict these intervals directly instead of categorizing ECGs into normal and abnormal groups. The predicted intervals and amplitudes include the PR interval, QRS duration, heart rate, J-point elevation, QT interval, R-wave amplitude, and T-wave amplitude (see supplemental figure S1). By predicting these measurements directly with regression, we allow using these intervals for better interpretation of the results rather than limiting it to a predefined set of categories. The second case study looks at differentiating between male and female ECGs.

All models are trained and evaluated on either raw 10-second 12-lead ECGs or on the 12SL-generated median beat from the GESUS dataset^22^. The performance of all GESUS generated models is replicated in the Inter99 dataset^23^. The demographics of the study populations are summarized in Table 1.

**Table 1.** Characteristics of the participants in both population studies. Values are presented as median [fifth to ninety-fifth percentiles] for continuous measures and % (*n*) for categorical variables.

| Variable | GESUS (training) | Inter99 (replication) |
| --- | --- | --- |
| Number of samples (*n*) | 8,939 | 6,667 |
| Age (years) | 56.6 [35.1; 78.4] | 45.3 [34.5; 60.0] |
| Female sex | 54.3% (4,852) | 51.2% (3,412) |
| BMI (kg/m<sup>2</sup>) | 26.1 [20.4; 35.3] | 25.7 [20.2; 34.8] |
| Heart rate (bpm) | 64 [48; 85] | 66 [51; 86] |
| QT interval (ms) | 408 [364; 460] | 402 [362; 450] |
| PR interval (ms) | 158 [126; 204] | 156 [124; 196] |
| QRS duration (ms) | 92 [76; 118] | 90 [76; 110] |
| J-point elevation V5 (μV) | -5 [-54; 48] | 4 [-35; 58] |
| R-peak amplitude V5 (μV) | 1,376 [698; 2,426] | 1,171 [600; 2,044] |
| T-wave amplitude V5 (μV) | 346 [122; 698] | 327 [122; 649] |

### CNN results

The performance of our method for the first use case, ECG intervals and amplitudes, is evaluated using quantitative regression metrics as seen in Table 2. The primary metrics used for evaluation are the mean absolute error (MAE) as it is easily interpretable, and the root-mean-squared error (RMSE) as it is more sensitive to outliers. In Table 2, we see that every model improves the ZeroR-estimate by a large margin. This shows that the proposed architecture successfully analyzes the ECG, both in the voltage and time domains. For interval measurements, the MAEs are close to two samples (4 ms) for both the median beat and rhythm strip (10 s) measurements. Amplitude measurements varied similarly (the least significant bit =4.88 µV), indicating that the network performed equally well with voltage and time-domain measurements.

**Table 2.** Training and validation error in GESUS^22^ and replication error in Inter99^2^

| Type | Variable | Validation on GESUS<br>(5-fold) |  | Replication on Inter99 |  | Zero R on Inter99 |  |
| --- | --- | --- | --- | --- | --- | --- | --- |
|  |  | MAE | RMSE | MAE | RMSE | MAE | RMSE |
| Median | QT Interval (ms) | 3.26 ± 0.80 | 5.08±0.40 | 3.13±0.19 | 4.89±0.19 | 21.7 | 27.4 |
|  | PR Interval (ms) | 2.82±0.15 | 4.52±0.49 | 2.73±0.06 | 4.70±0.23 | 17.6 | 22.8 |
|  | QRS duration (ms) | 2.98±0.15 | 4.10±0.22 | 2.58±0.07 | 3.43±0.07 | 9.0 | 11.6 |
|  | Heart Rate (beats per min) | 1.54±0.07 | 2.44±0.09 | 1.57±0.06 | 2.33±0.07 | 8.6 | 11.1 |
|  | J-point elevation (µV) | 8.16±0.40 | 11.20±0.69 | 5.77±0.10 | 8.09±0.12 | 22.2 | 29.0 |
|  | T-wave amplitude (µV) | 5.63±1.31 | 15.2±6.83 | 5.80±1.13 | 16.10±0.53 | 129.0 | 167.0 |
|  | R-peak amplitude (µV) | 8.60±1.05 | 16.00±4.13 | 8.30±0.98 | 21.70±0.71 | 413.0 | 501.0 |
| Rhythm | QT Interval (ms) | 3.97±0.03 | 6.05±0.39 | 3.62±0.03 | 5.82±0.20 | 21.7 | 27.4 |
|  | PR Interval (ms) | 3.67±0.21 | 5.60±0.60 | 3.58±0.60 | 5.80±0.31 | 17.6 | 22.8 |
|  | QRS duration (ms) | 3.08±0.12 | 4.33±0.17 | 3.39±0.06 | 4.49±0.07 | 9.0 | 11.6 |
|  | Heart Rate (beats per min) | 0.31±0.01 | 0.40±0.02 | 0.18±0.01 | 0.6±0.1 | 8.6 | 11.1 |
|  | J-point elevation (µV) | 10.50±0.31 | 14.10±0.50 | 7.90±0.19 | 10.70±0.24 | 22.2 | 29.0 |
|  | T-wave amplitude (µV) | 11.50±0.43 | 25.00±5.55 | 9.40±0.41 | 19.40±1.79 | 129.0 | 167.0 |
|  | R-peak amplitude (µV) | 20.10±0.70 | 33.00±4.54 | 17.4±0.55 | 51.00±13.21 | 413.0 | 501.0 |

### Attention maps

To be able to create meaningful and detailed visualizations, we modified the GradCAM approach so that visualizations are generated for each lead of the ECG, where the final attention maps are produced by averaging the importance values across all leads. We call this method ECGradCAM since it is able to give a more accurate representation of what regions of the ECG are most important for the model. We focus our interpretation on the last layer of the last residual module of the neural network (as shown in supplemental figure S2). This corresponds to the final layer before prediction, meaning the visualizations show what areas of the ECG are deemed most important at the moment of prediction. It can also be useful to interpret the intermediate layers of a network^25^ as these layers may offer insight into how the network’s perception changes and how it narrows down the analysis to the final result (see supplemental figure S3). Here, we note that the initial residual block recognizes several features in the ECG, which becomes more and more focused on the relevant wave as the ECG progresses through the residual layers.

The attention maps often highlight the areas we expect when predicting a specific interval or amplitude. Figure 1 presents a median ECG visualized for six of the predicted variables. For instance, the QRS complex is highlighted when we predict QRS duration, and the end of the T-wave is delineated along with the beginning of the QRS complex for QT interval measurement. For amplitude measurements, the corresponding wave top is correctly pinpointed by the attention maps. One should note that for amplitude measurements, other parts of the ECG are given minor importance, likely for the network to learn about the ECG voltage baseline. For intervals, secondary activations are also observed, such as the T-wave being highlighted when measuring the PR-interval. We hypothesize that these secondary activations may be happening because the network needs to appreciate the whole ECG in order to narrow its search down and perform the actual measurements. This is further supported by the PR interval attention maps generated for the intermediate layers (see supplemental figure S3), where the network highlights the QT in the former layers, but less so at the moment of prediction.

**Figure 1.**
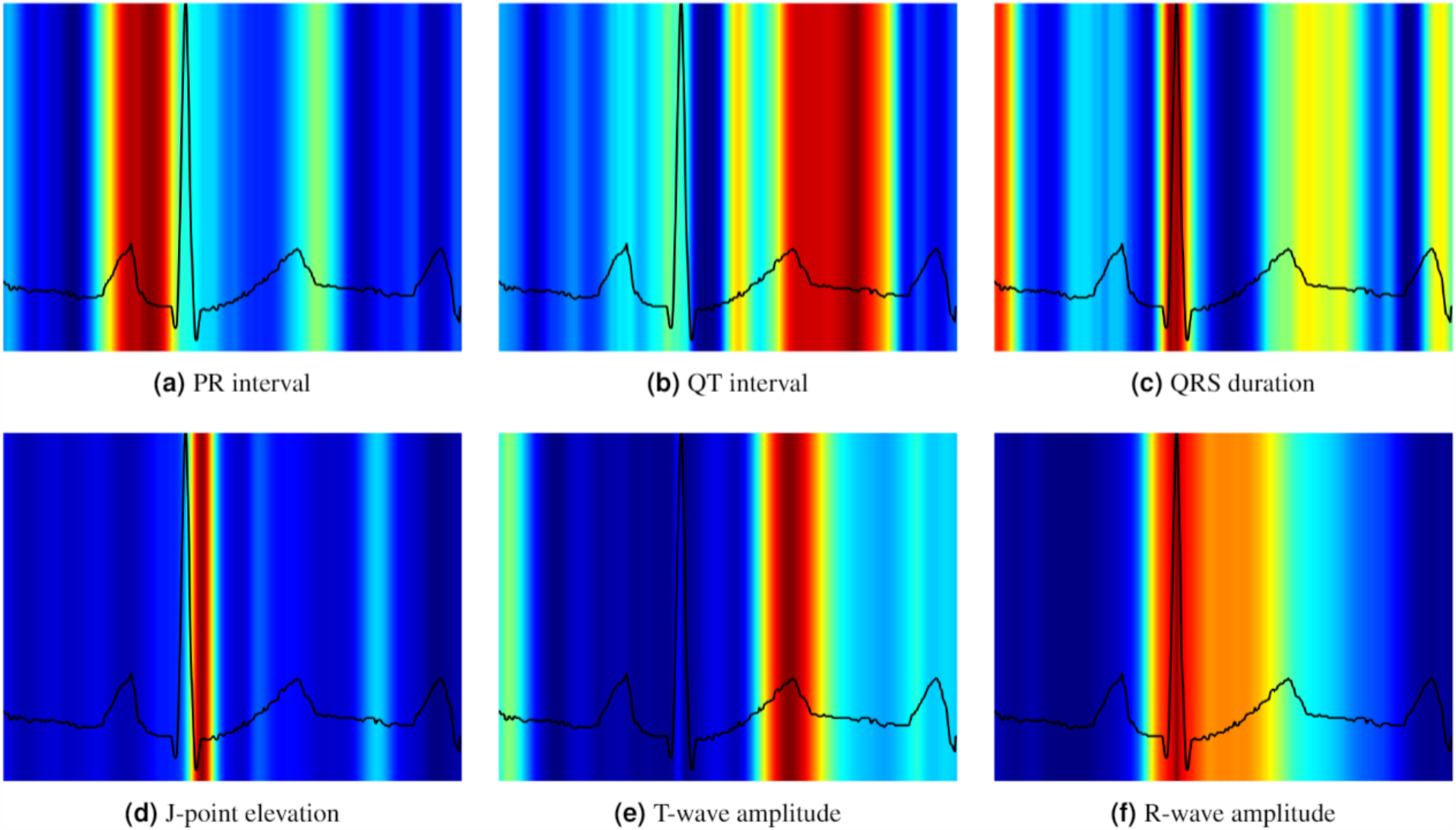
Visualization of the attention maps generated for the interval and amplitude prediction models. As we can see from the plots, the model learns to inspect the waves and intervals that are related to the predicted variable. Red color indicates high importance and blue color low importance of the ECG for the decision of the neural network.

### Sex prediction

For a cardiologist, the task of determining a subject’s sex from the ECG is nearly impossible. However, our network is able to correctly identify the sex with an accuracy of 89% (Table 3). Here, we can see the potential of attention maps, as the accuracy output from the network does not give any clue or insight into how the network made its decision on sex classification. The attention maps (see Figure 2) clearly show that the ECG sex classification is mainly based on the QRS complex and more specifically on the downslope of the R-wave, offering new insight into electrophysiology. Using findings from the attention maps, we did a post-hoc analysis with logistic regression predicting sex using QRS duration, R- and S-amplitudes and the timing of the R- and S-waves. It revealed an accuracy of 73% (our CNN: 89% QRS duration alone: 69%) and an AUC of 0.80 (our CNN 0.96 QRS duration alone: 0.72). The wave blocking experiments, which can be found in the methods section, verified this observation, since removing the P-wave has nearly no influence on the accuracy of the sex prediction and removing the T-wave had only minor influence, whereas removing the QRS wave resulted in drastic reduction in performance. This shows that one can obtain new knowledge by using our ECGradCam method in combination with the deep neural network.

**Table 3.** Mean absolute error ± standard deviation measured on the replication dataset when blanking specific waves of a median heartbeat. Prediction errors also increased dramatically when the feature in question is blanked out. Prediction errors also often increased slightly when other parts of the ECG are blanked.

| Variable | Median ECG | Blanking P-Wave | Blanking QRS complex | Blanking T-Wave |
| --- | --- | --- | --- | --- |
| QT interval (ms) | 3.10±0.20 | 3.20±0.20 | 32.00±6.40 | 47.00±11.00 |
| PR interval (ms) | 2.70±0.10 | 33.00±9.00 | 41.00±5.20 | 3.80±0.60 |
| QRS duration (ms) | 2.60±0.10 | 4.00±0.40 | 41.00±5.20 | 3.50±0.10 |
| Heart Rate (bpm) | 1.57±0.06 | 2.92±0.17 | 3.62±1.66 | 4.79±0.97 |
| J-point elevation ($\mu V$) | 5.80±0.20 | 6.40±0.50 | 23.00±2.90 | 8.60±0.40 |
| T-wave amplitude ($\mu V$) | 5.80±1.10 | 6.10±1.30 | 8.70±1.30 | 339.00±7.00 |
| R-wave amplitude ($\mu V$) | 8.40±1.00 | 8.60±1.00 | 927.00±14.00 | 10.50±3.10 |
| Sex classification (%) | 88.80±0.70 | 87.50±1.00 | 62.40±6.60 | 79.80±2.50 |

**Figure 2.**
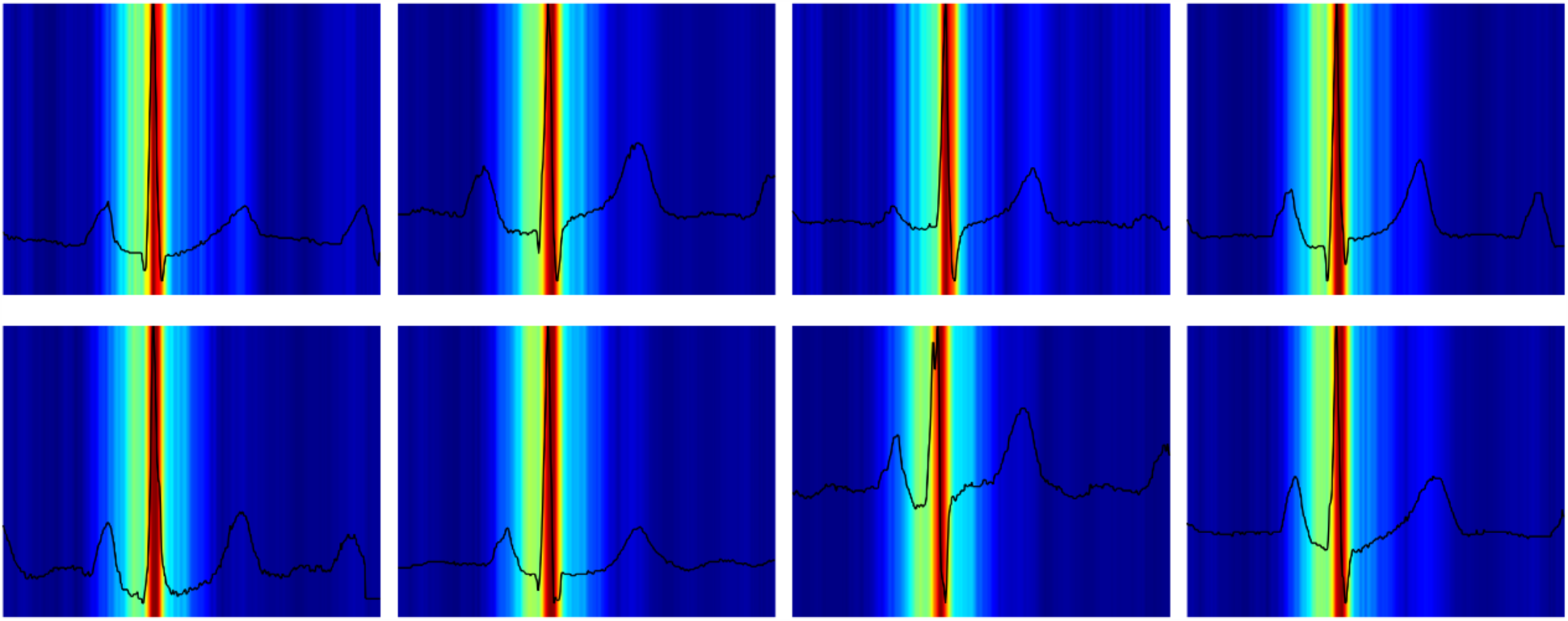
Visualizations of the attention maps from the sex classification model for eight different ECGs. The plots suggest that the QRS-complex and especially the downslope of the R-wave is most important when distinguishing between a male and female ECG. Red color indicates high importance and blue color low importance of the ECG for the decision of the neural network.

### Human Cardiologist vs neural network evaluation

To assess how the neural network compares to standard clinical decision making, we further evaluate the performance of our model by comparing its predictions to predictions made by cardiologists who have manually annotated a set of twenty randomly selected ECGs from the Inter99 replication dataset. As seen in Table 4, the trained networks prove substantially more precise and consistent than human expert assessments. Human bias-corrected MAE and RMSE are around 15-20 milliseconds, i.e., a factor 4-5x higher than the neural network. Errors in heart rate measurements are below 1 beat per minute (BPM) for the network, but about 3 BPM for the human operators with multiple errors above 10 BPM. Amplitude measurements are way more difficult for humans, given the resolution of the ECG and the accuracy of the human eye. For the R-peak amplitude, the network operates at a MAE of twice the least significant bit at 4.8 microvolts. In contrast, the human reviewer operates with a MAE in the magnitude of half a millimeter (corresponding to 50 microvolts 10x least significant bit).

**Table 4.** Evaluation of twenty randomly selected ECGs by two experienced cardiologists. Bias is the average difference between the ground truth and the doctor/network. Mean absolute error (MAE*) is the bias-subtracted mean absolute error, to account for the fact that there is no universal correct measurement for an ECG, ensuring that any personal bias does not contribute to the error (e.g., some doctors measure QT consistently shorter or longer than others). R: Pearson’s correlation coefficient between the doctor’s two measurements/the neural networks five folds. SD: Standard deviation.

| Variables | Test set,<br>mean±SD | Neural Network |  |  | Doctor A |  |  | Doctor B |  |  |
| --- | --- | --- | --- | --- | --- | --- | --- | --- | --- | --- |
|  |  | Bias | MAE* | R | Bias | MAE* | R | Bias | MAE* | R |
| Heart rate (bpm) | 71 ± 8 | 0.04 | 0.20 | 0.99 | 0.98 | 3.20 | 0.68 | -0.52 | 1.38 | 0.99 |
| QT interval (ms) | 392 ± 21 | 0.50 | 3.70 | 0.98 | -8.50 | 15.8 | 0.49 | -17.8 | 9.2 | 0.89 |
| QRS duration (ms) | 91 ± 10 | -3.30 | 3.00 | 0.98 | -7.80 | 11.9 | 0.39 | -7.6 | 8.22 | 0.55 |
| PR interval (ms) | 161 ± 16 | -2.50 | 4.70 | 0.99 | 5.90 | 8.00 | 0.87 | 6.45 | 9.01 | 0.82 |
| R-peak amplitude<br>(mm) | 12 ± 4 | 0.02 | 0.16 | 0.99 | 0.57 | 0.42 | 0.98 | 0.06 | 0.50 | 0.95 |
| J-point elevation<br>(mm) | -0.04 ± 0.28 | 0.02 | 0.09 | 0.98 | 0.12 | 0.19 | -0.01 | -0.01 | 0.17 | 0.86 |
| T-wave amplitude<br>(mm) | 3.00 ± 1.50 | 0.03 | 0.10 | 0.99 | -0.13 | 0.32 | 0.89 | 0.20 | 0.56 | 0.80 |

## Discussion

The present paper identified three novel findings. First, we present a residual CNN that reliably analyzes both ECG intervals (time dimension) and amplitudes (voltage dimension) independent of whether the ECG presented is a 10-second 12-lead ECG or a 1.2-second median representative beat. The architecture proved accurate for a variety of different ECG tasks. In all cases examined, algorithmic prediction outperformed the cardiologists by a large margin. Furthermore, with repeated blinded measurements, the cardiologists had a large intra-observer variation, whereas the neural network is very consistent in its predictions between folds. The MAE is between 3-4 ms corresponding to two samples which is close to the physical obtainable lower limit (since the intervals consist of two cumulative uncertainties, one in the beginning and one at the end of the interval). In general, measurements are more accurate when using 1.2-second median beats compared to 10-second rhythm strips, except for heart rate. The generation of the median beat reduces noise by averaging all beats during the 10-seconds ECG, stretching each complex to minimize the influence of variations, and therefore being more accurate. The exception of heart rate is not surprising since several ECG complexes are needed to properly estimate heart rate, and these are only found in the rhythm strip. More surprising is the finding that the neural network, by using other features than the RR-interval to calculate heart rate, is able to obtain a relatively good estimate of heart rate from the single ECG complex of the median beat. In fact, the heart rate estimate from the median is only slightly worse than that based on the rhythm strip (see Table 2). Second, we presented ECGradCAM attention maps for 12-lead ECG-analysis to explain how the network made its decision. In medical practice, explainability is crucial because medical doctors are concerned that algorithms may produce erroneous results, either due to bias or trying to predict outcomes not appropriately represented in the training data. For example, measurement of the PR interval in a case of ventricular tachycardia would be unreliable if the training set only consisted of ECGs taken in sinus rhythm. Interpretation and transparency should be at the forefront when developing new algorithms intended for medical use. Although the results suggest that deep learning could be an essential tool for cardiologists doing analysis and interpretation of ECG, it is doubtful that the neural network models alone would be accepted by doctors. By themselves, the presented models without the accompanying attention maps or obscurity visualization are not transparent and left little to no information for interpretation besides the output produced.

The attention maps showed nicely that all amplitude measurements focused on the proper ECG wave, and in cases of interval measurements, both the beginning and the end of the specific interval are highlighted by the algorithm. One may notice that the beginning or end of the interval is not necessarily bright red in the attention maps, which can be seen with the QT interval in Figure 1b. The ECGradCAM technique shows the importance of the different parts in the ECG at the level of the last convolutional layer. As long as the relevant information is fed to the fully connected layer, the fully connected layer may amplify or attenuate the signal as needed to give the best performance. When the attention map gives sensible explanations for the network’s decision, it will open up for more acceptability and trust among the medical doctors who use the neural network for ECG analysis. The obscurity tests confirm the results given by the attention map. When ECG waves used for the specific interval or amplitude measurement are removed, the MAE increased dramatically, confirming the message from the attention maps that our trained neural network focused on the same features as human cardiologists, just more accurately and reliably. One may also notice that the network also tries to extract information about the baseline from the ECG. Since we make use of batch normalization (a standard feature in neural networks to avoid exploding gradients), the network had to get an idea of the magnitude of normalization to restore the absolute values needed for amplitude prediction. This may be the reason why the network also wanted attention on more steady, constant parts of the ECG. By providing these explanations with a predicted variable, we allow the users to interpret the results with confidence that model had some notion of the traits that make up the variable in question.

The sex prediction is an excellent example how neural networks in combination with an explanation method can be used to discover novel medical knowledge. It is well known that there are sex differences in the ECG. Female ECGs on average have a longer QT time, faster heart rate, and shorter QRS duration^26^. However, if one would ask a cardiologist to determine sex based on an ECG alone, they would not be able to make a confident prediction. Recent studies using deep learning have shown that neural networks can differentiate between the sexes from the ECG alone, but the underlying reasoning is not provided. The attention maps indicate that the physiological background seems to be differences in the R-wave downslope, which may provide important mechanistic insight into the observed sex differences. We confirmed the findings by our neural network, that simple logistic regression with: QRS duration, wave amplitudes and timing (slopes can be inferred by wave amplitudes and timings) significantly improved sex prediction compared to QRS duration alone. Although adding the R-wave downslope to the QRS duration significantly increased sex prediction, the neural network clearly still performed better than the logistic regression. This either indicates that nonlinear information in the R-wave downslope may be important or that only part of the downslope predicts the sex. These results are a scholarly example that the use of attention maps can assist the scientist in getting new insight and identify novel hitherto unknown features not only for classification but important for physiological understanding. Classifying an appropriate outcome in a suitable population, one may identify novel prognostic markers in the ECG for that outcome, which may lead to a suggestion for possible treatments.

However, why do these networks work so well for ECGs, and what features are analyzed to make such accurate predictions? These questions are common in machine learning, where we often have high-performance algorithms that are difficult to understand. In medicine, this is a known challenge as medical doctors seek evidence and require more than a numerical output from a system to make a decision, ensure good quality, and avoid automation bias^27,28^. Given the two previously tasks, we show that attention maps can offer sensible explanations for predicting key characteristics of an ECG, and that they can also discover new features previously unknown to the medical community. Not only does this help us better understand the “thought process” behind the algorithm’s predictive framework, but it also opens the possibility to create increased trust and acceptance among the medical doctors who would use the system. Deep learning could be an essential tool for cardiologists doing analysis and interpretation of ECG going beyond simple automation tasks. By themselves, the presented models have little to no transparency and leave little to interpret besides the produced output. Therefore, the use of attention maps to aid in the interpretation of model predictions as presented here is essential if these types of algorithms are to be trusted by the medical community. Interpretation and transparency should be at the forefront when developing new algorithms intended for medical use.

## Conclusions

In this paper, we present a study on interpreting deep learning models for ECG analysis. We introduce a neural network architecture which predicts multiple attributes of a standard median or 10-second rhythm ECG with high accuracy. The model was compared against real-world cardiologists, where our model the cardiologists by a large margin. The predictions are interpreted using attention maps (ECGradCAM), which show how the network operates and confirmed that the neural network analyzes ECGs in a similar manner to that of cardiologists. Furthermore, we show that the neural network can differentiate between male/female ECGs with over 90% accuracy. The ECGradCAM attention maps reveal that the down-slope in the QRS-complex is the most important feature of an ECG when determining sex.

## Methods

### Data Populations

We use digital ECGs from two population studies. 1) The Danish General Suburban Population Study (GESUS)^22^ consisting of 8,939 free-living subjects (age 56.5±13.5, 54% females) at least 18 years old from the Naestved municipality, 90 km south of Copenhagen, the Capital of Denmark, randomly chosen. The study was approved by the local ethics committee (SJ-113, SJ-114, SJ-147, SJ-278). 2) The Inter99 study (CT00289237, ClinicalTrials.gov) consists of 6,667 free-living subjects (age 46.1±7.9, 51% females) randomly drawn from the Glostrup municipality with an age of 30-65 years^23^. This yields a collection of ECGs from people with and without cardiac disease, and an equal representation of men and women. Both studies are conducted in accordance with the Declaration of Helsinki.

### Electrocardiography

All ECGs are digitally recorded as 10-second ECGs with 12 leads. All ECGs are transferred to a MUSE Cardiology Information system (GE Healthcare, Wauwatosa, WI, USA) and ground truths are calculated with version 21 of the Marquette 12 SL algorithm (GE Healthcare, Wauwatosa, WI, USA). The ECGs are recorded with a sample rate of 500 Hz and a resolution of 4.88 microvolts per least significant bit.

### Prediction Model

#### Architecture

A digital electronic ECG can be represented as a two-dimensional matrix of integers representing voltage at a specific point in time. To analyze these measurements, we use a standard convolutional neural network (CNN) consisting of eight residual modules (as introduced by He et al.^21^) to capture the complex features and relationships present in a standard ECG. The neural network architecture is built to handle two different types of input, a single representative median heartbeat of 1.2-second duration and a 10-second rhythm ECG. Both input types contain data from 12-lead ECGs. A detailed view of the neural network architecture can be seen in supplemental figure S2. From the input layer, the ECG is passed through two convolutional layers before being average pooled. The two convolutional layers generate 64 and 32 feature maps with a kernel size of 8 and 3. After this initial convolution block, the output is sent through eight residual blocks, each consisting of two convolutions. Each convolutional layer in the residual blocks generates 64 and 32 feature maps, respectively, and both layers use a kernel size of 50. We use a large kernel size to extend the receptive field to include multiple parts of a typical ECG. This could, for example, capture both the P wave and the QRS complex in a single convolution. We add batch normalization after each convolution and dropout after the final convolution with a drop-rate of 50%. After the eight residual blocks, the output is globally average pooled before making the final prediction. The prediction layer consists of a single neuron with a linear activation that predicts a single variable of the ECG.

#### Training

All models are trained for a maximum of 1,000 epochs on a computer consisting of two Intel Xeon Silver 4116 CPUs running at 2.1GHz, four Nvidia RTX 2080Ti graphics cards, and 96 gigabytes of RAM. The models are implemented using Keras version 2.1.0 with a TensorFlow backend on Ubuntu 18.04.2. To optimize the weights, we used the gradient descent-based Nadam^29^ with a learning rate of 0.0005. The learning rate is selected based on manual testing and prior experience from our previous works^25^, otherwise, we used the Keras defaults for all optimizer parameters. In total, we performed 14 different experiments, seven using the median for prediction and seven using the rhythm. The variables predicted with regression parameters (include the QT interval, PR interval, QRS duration, heart rate, ST-segment deviation from baseline at the junction (STJ), T-wave amplitude, and R-peak amplitude). The three amplitudes are lead specific and lead V5 is used. Some of these variables cater more to rhythm analyses (such as heart rate), while others are more appropriate for median complexes (such as the R-peak). One problem with training on the median complexes is that they are all centered in a manufacturer-specific way, whereby each wave appears in nearly the same place in each of the ECG. Thereby, the network can learn to predict a particular vicinity and guarantee a relatively low error. To circumvent this problem, we time-shifted all median complexes by a random amount (−40 to +40 milliseconds) so that the network learns to find the individual waves. This increases the likelihood that the model can be used on ECGs from other manufactures with different temporal alignment. No alignment is performed for the rhythm ECGs; the start of the recording is random with respect to the ECG. Furthermore, to test the networks ability to classify in binary outcomes, we classified the ECG for sex (male/female).

#### Attention maps

To obtain physiological insights from the neural network’s decisions, it is necessary to understand how and why a decision is achieved. In this study, we used attention maps to visualize which parts of the ECG have importance for each interval/amplitude prediction. To explain the predictions of our model, we use gradient-based activation maps (attention maps) to visualize which parts of the ECG are the most important when predicting a given variable or class. The technique used is a modified version of GradCAM^24^, commonly used to interpret image classification models. As we show in our study, this approach works just as well for regression tasks of quantitative measurements in the data. Visualizations are generated based on a given network layer and output neuron, and produce a heat map, which marks the most important areas as hot (red color) and the less important regions as cold (blue color). In this context, importance signifies how much weight a specific area contributes to the overall prediction. We are not the first to use attention maps to interpret deep neural networks applied to ECGs^18,19^. Most other works use these visualizations to confirm that their model does not deviate from the expectations of the medical doctors^20^. Our work goes one step further and expands the method of explanations to find new insights into the unique properties of ECGs through a case-study on sex classification. Furthermore, even though the attention maps are generated on a per-lead basis, we average the explanations for each lead to produce an explanation that contains more fine-grained details and thus is able to more accurately represent what regions of the ECG are most important for the model when making a specific prediction.

#### Evaluation

To ensure a fair and robust evaluation, we trained each model with five-fold cross-validation for 1,000 epochs on the GESUS dataset^22^, resulting in 7,152 samples being used for training and 1,787 for validation. After training and internal cross-validation, the results of GESUS models are replicated in the Inter99 dataset^23^ to examine whether the models are generalizable or not. As seen in Table 1, GESUS and Inter99 datasets are comparable regarding ECG measurements, although participants in the GESUS study are on average older than participants in the Inter99 study. The neural network performance is evaluated by the MAEs (|predicted – actual value|) and the 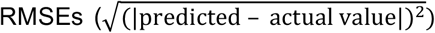 to evaluate the mistakes of the neural networks relative to the ground truth. To give an idea of the magnitude of uncertainty, we calculated the ZeroR-estimate, defined as constantly guessing the population mean of the desired variable. If a model’s performance is not better than ZeroR, the model has not learned anything except the population mean. Conversely, if the model performance is better than the ZeroR, it follows that the model has succeeded in extracting and processing features from the ECGs.

Furthermore, ECGs were evaluated manually by two skilled cardiologists. Whereas the neural network by definition has a bias (i.e., average error) of zero (ignoring an eventual bias in the ground truth from the 12SL algorithm), the human overreaders may exhibit substantial bias relative to the ground truth (i.e., the measure consistently shorter or longer intervals), which originates from their own training and personal preference. Since this bias is not an error, the human bias is subtracted from the errors before the calculation of human MAE and RMSE.

### Wave blocking

To verify that the neural network model is focusing on the relevant waves of the ECG, and as an alternative to the attention maps, we remove specific parts of the ECG (either the P-QRS- or T-wave) from the median ECGs of the replication set. Using the MUSE 12SL fiducial points, we blank out a wave by replacing it from the start to the end with a lead-specific linear interpolation. This analysis represents an alternative measure of explainability for representative beats of an ECG by analyzing the decrease in performance when different waves of the ECG are blanked. Thus, we can test how dependent our model is on different parts of the ECG and verify which waves the model is focusing on when making a prediction. The results in Table 3 show that the model performance drop when the wave involved with a particular feature is removed. However, we also find that removing non-involved waves typically decreases performance slightly, suggesting that the neural network also includes other parts of the ECG to stabilize the model to ensure that it is analyzing the correct part of the ECG.

## Data Availability

The data is not available to the public.

## Data Availability

The data is not available to the public.

## Code Availability

The code used to conduct the experiments and generated the related attention maps is available on GitHub at https://github.com/Stevenah/ecg-attention-maps.

## Acknowledgements

This work is funded in part by Novo Nordisk Foundation project number NNF18CC0034900.

## Author contributions

Steven A. Hicks, Jonas L. Isaksen, Vajira Thambawita, Michael A. Riegler, and Jørgen K. Kanters conceived the experiment(s).

Steven A. Hicks, Jonas L. Isaksen, and Jørgen K. Kanters conducted the experiment(s).

Steven A. Hicks, Jonas L. Isaksen, Vajira Thambawita, Michael A. Riegler, and Jørgen K. Kanters analyzed the results.

All authors reviewed the manuscript.

## Competing Interests statement

### The authors declare the following competing interests

None

### The authors declare no competing interests

Steven A. Hicks, Jonas L. Isaksen, Vajira Thambawita, Jonas Ghouse, Gustav Ahlberg, Allan Linneberg, Niels Grarup, Inga Strümke, Christina Ellervik, Morten Salling Olesen, Torben Hansen, Claus Graff, Niels-Henrik Holstein-Rathlou, Pål Halvorsen, Mary M. Maleckar, Michael A. Riegler, and Jørgen K. Kanters.

