## Supplemental Figure 1 for "Explaining Deep Neural Networks for Knowledge Discovery in Electrocardiogram Analysis"

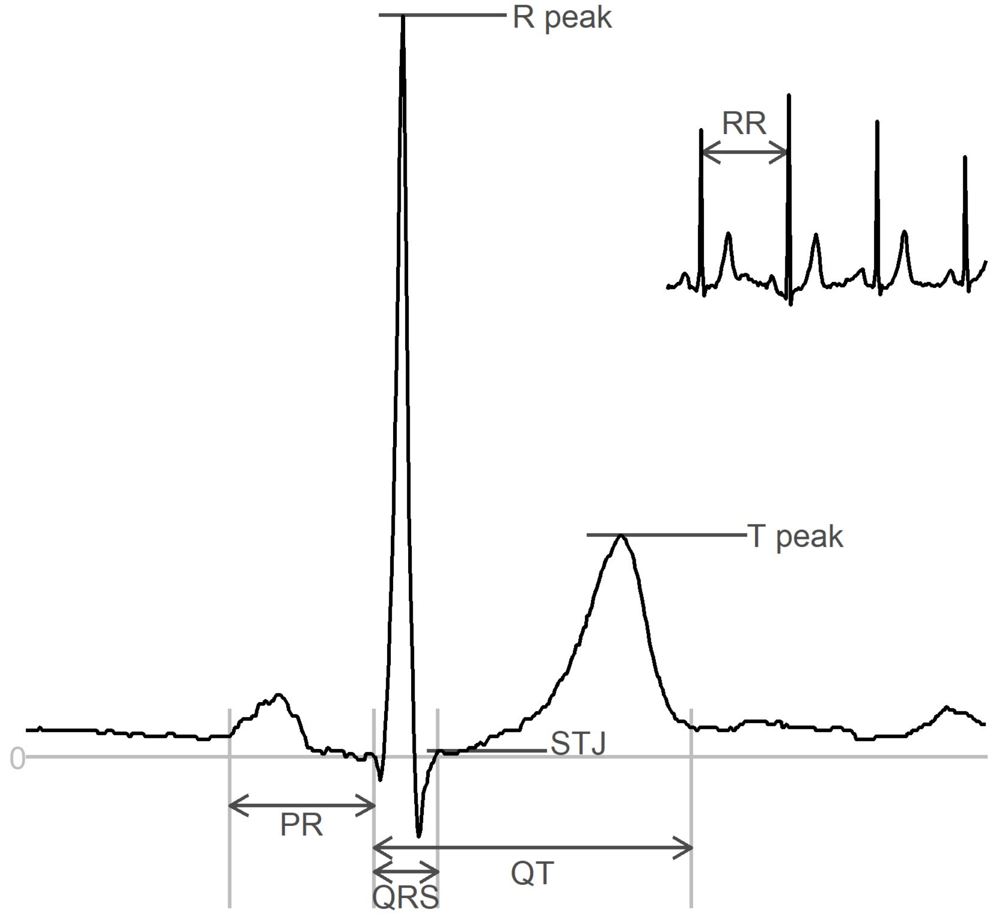


**Figure S1.** An annotated ECG representative beat and rhythm strip (top-right inset) with intervals (PR, QRS, QT) and amplitudes (Rpeak, STJ, Tpeak). Amplitudes are measured with respect to the baseline. STJ denotes J-point elevation. Heart rate is calculated as HR = 60,000/RR where RR is measured in milliseconds.
