## Supplementary figures and images for "Explaining Deep Neural Networks for Knowledge Discovery in Electrocardiogram Analysis"

### Supplemental Figure 2

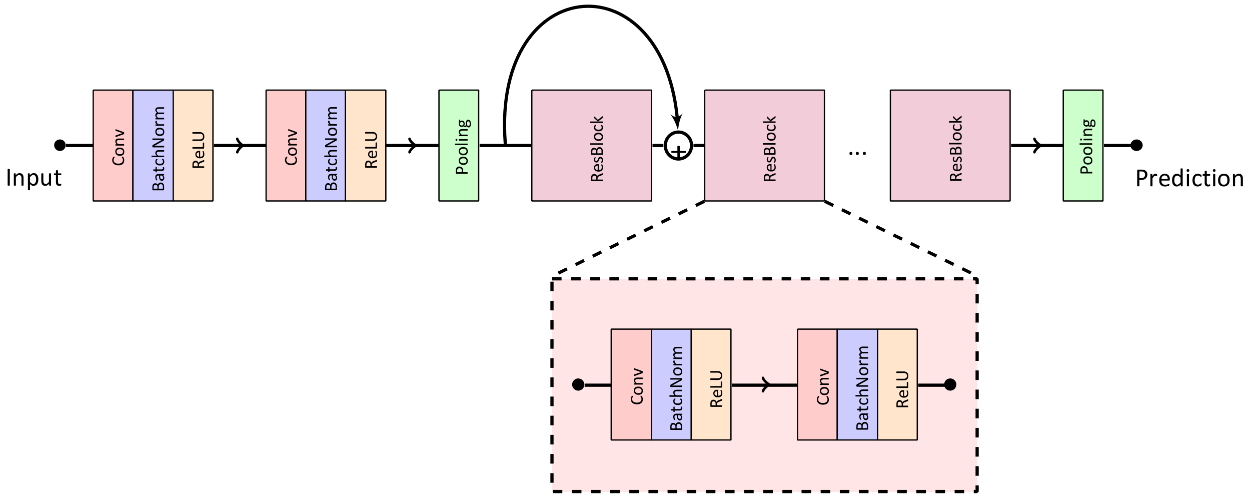


**Figure S2.** The convolutional neural network-based architecture used for all experiments.
