## Supplemental Figure 3 for "Explaining Deep Neural Networks for Knowledge Discovery in Electrocardiogram Analysis"

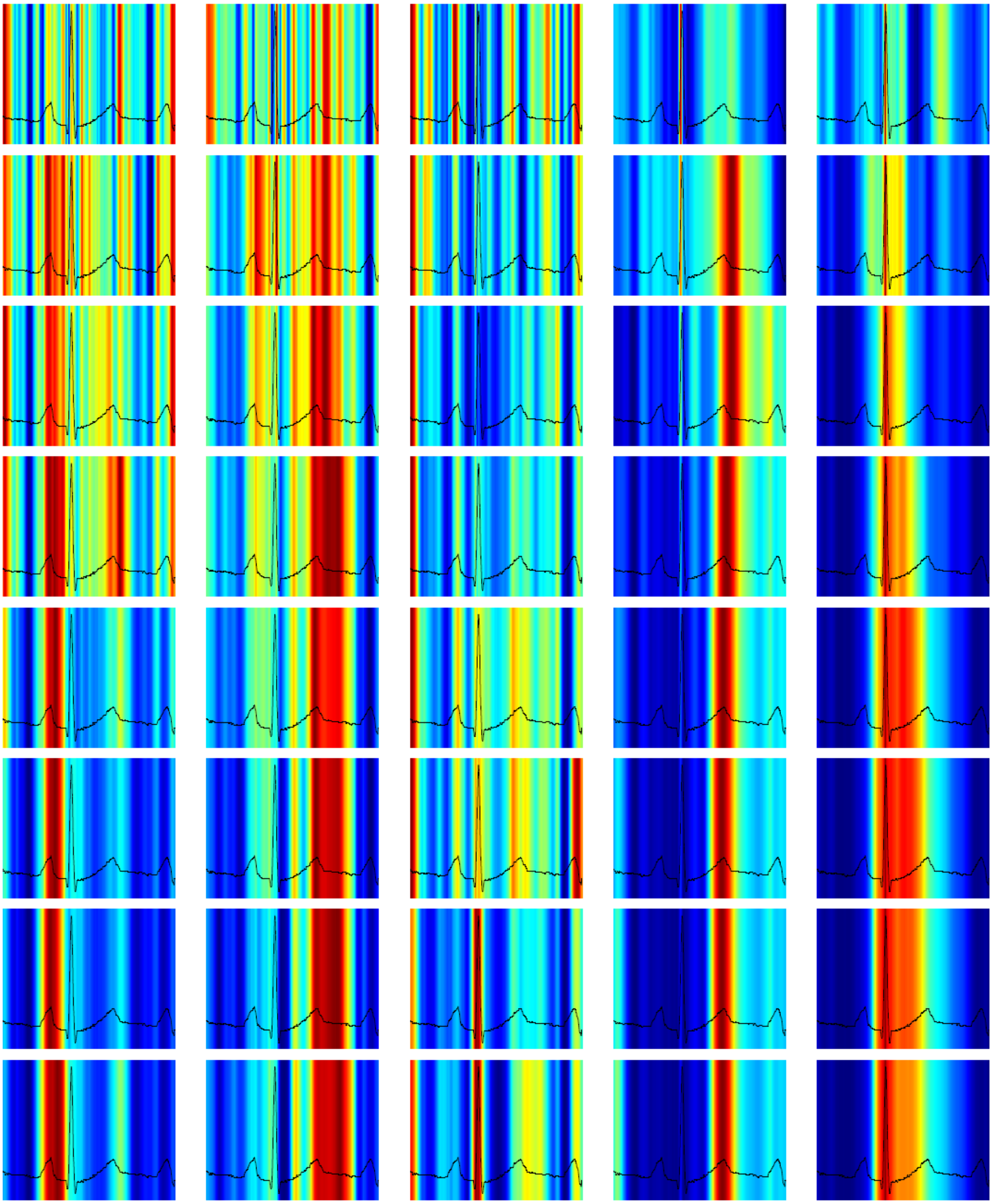

(a) PR interval (b) QT interval (c) QRS complex (d) T-wave amplitude (e) R-wave amplitude

**Figure S3.** Attention maps from intermediate convolutions. First row is last convolution in the first residual block, second row is last convolution in second residual block etc.
